# Glucose-6-Phosphate Dehydrogenase (G6PD) Deficiency and Long-Term Risk of Immune-Related diseases

**DOI:** 10.1101/2023.03.23.23287616

**Authors:** Ariel Israel, Alejandro A Schäffer, Matitiahu Berkovitch, David J. Ozeri, Eugene Merzon, Ilan Green, Avivit Golan-Cohen, Eytan Ruppin, Shlomo Vinker, Eli Magen

## Abstract

**BACKGROUND:** Glucose-6-phosphate dehydrogenase (G6PD) deficiency is an X-linked recessive enzymatic disorder, particularly prevalent in Africa, Asia and the Middle East. In the US, about 14% of black men are affected. Individuals with G6PD deficiency are often asymptomatic but may develop hemolysis following an infection or upon consumption of specific medications. Despite some evidence that G6PD deficiency affects the immune system, the long-term health risks associated with G6PD deficiency had not been studied in a large population.

**METHODS:** In this retrospective cohort study, health records from G6PD deficient individuals were compared to matched controls in a national healthcare provider in Israel (Leumit Health Services). Rates of infectious diseases, allergic conditions and autoimmune disorders were compared between groups.

**RESULTS:** The cohort included 7,473 G6PD deficient subjects (68.7% men) matched with 29,892 control subjects (4:1 ratio) of the same age, gender, socioeconomic status and ethnic group, followed during 14.3±6.2 years.

Significantly increased rates for autoimmune disorders, infectious diseases and allergic conditions were observed throughout this period. Notable increases were observed for rheumatoid arthritis (OR 2.41, p<0.001), systemic lupus erythematosus (OR 4.56, p<0.001), scleroderma (OR 6.87, p<0.001), pernicious anemia (OR=18.70, P<0.001), fibromyalgia (OR 1.98, p<0.001), Graves’ disease (OR 1.46, P=0.001), and Hashimoto’s thyroiditis (OR 1.26, P=0.001). These findings were corroborated with elevated rates of positive autoimmune serology and higher rates of treatment with medications commonly used to treat autoimmune conditions in the G6PD deficient group.

**CONCLUSION:** G6PD deficient individuals suffer from higher rates of autoimmune disorders, infectious diseases, and allergic conditions.

## Introduction

Glucose-6-phosphate dehydrogenase (G6PD) deficiency is the most common enzymatic X-linked human disorder and it primarily affects red blood cells (Cappellini and Fiorelli, 2008). An estimated 400 million people worldwide carry a mutation in the G6PD gene associated with enzyme deficiency, with marked ethnic and geographic differences (Luzzatto et al., 2016; Nkhoma et al., 2009). G6PD deficiency is prevalent in Africa, Asia and the Middle East. In the United States, about 14% of African-American men are affected (Ruwende et al., 1995; Tishkoff et al., 2001). There are numerous alleles of different severities and geographic propensities (Beutler, 1994). Most individuals with G6PD deficiency remain asymptomatic or experience mild symptoms throughout their lives but may develop hemolysis under certain circumstances such as neonatal jaundice, consumption of specific medications, or following an infection.

Several studies have suggested that G6PD-deficient individuals may be more predisposed to specific autoimmune diseases (Dore et al., 2023; Saha et al., 1982), infections (Abu-Osba et al., 1989; Mondal et al., 2022; Rostami-Far et al., 2016) and diabetes mellitus (Lai et al., 2017; Saha, 1979). Associations were found between G6PD deficiency and cardiovascular risk (Cocco et al., 1998; Dore et al., 2021; Meloni et al., 2008; Pes et al., 2019) in conflicting directions. Regarding cancer, some studies found lower rate of cancer in G6PD deficient patients, while others found no difference (Cocco, 1987; Dore et al., 2016; Ferraris et al., 1988).

To date, there is no large-scale study showing the association of G6PD deficiency with infectious and immune-related diseases altogether. This large-scale observational study addresses this knowledge gap.

## Methods

### Cohort data source

We performed a retrospective cohort study using data from Leumit Health Services (LHS), one of the four nationwide healthcare providers in Israel(Israel et al., 2021). LHS has been using centrally managed electronic health records (EHR) for the last 20 years. The EHRs includes demographic data, physical measures, laboratory test results, medication prescription and purchases, and diagnosed conditions, documented by physicians according to International Classification of Diseases 9th Revision (ICD-9). The ethics committee of Leumit Health Services gave ethical approval for this work (LEU 02-23) with a waiver of informed consent.

Data were collected from the LHS data warehouse as of December 31, 2022. The study cohort was drawn from a pool of over 1,000,000 individuals who had been insured at least two years by LHS up to the date of data extraction.

### Cohort design

The study involves the comparison of two matched groups of individuals. The G6PD deficiency group consists of patients with a documented diagnosis of G6PD deficiency or a recorded laboratory test of G6PD activity performed in LHS, resulting in a measurement below 4 U/g Hg.

The control group was randomly selected from the pool of individuals without G6PD deficiency using an algorithm that requires exact matching for gender, age, geographic area based socio-economic status (SES) category, and ethnic group (General, Jewish Ultra-Orthodox, and Arabs), and year of first documented visit in LHS EHR system, to G6PD individuals, with a ratio of 4 controls for each included G6PD deficient patient. Individuals with G6PD deficiency the matching algorithm was not able to find the required number of control individuals exactly matching these variables were left out from the analysis.

Demographic records were extracted from LHS data warehouse, along with ICD-9 diagnosis records matching a list of pre-defined conditions, the last recorded body mass index (BMI), blood pressure (BP) measures, and smoking habits. The last laboratory test results for a set of predefined laboratory measures performed by LHS laboratory facilities were also extracted.

### Statistical Methods

Statistical association was assessed using Fisher’s Exact Test for binary variables, with odds-ratio and 95% confidence interval (CI). Numerical variables were tested using the Mann-Whitney U test, which does not assume a normal distribution. Cohort data extraction was performed using programs developed by Leumit Research Institute in Python. Statistics were computed in R statistical software, version 4.0.4.

## Results

### Demographic and laboratory characteristics

Among the pool of 1,031,354 LHS members with at least two years of medical history in the EHR, we identified 7,827 individuals with either a G6PD deficiency diagnosis or laboratory evidence of G6PD deficiency. The matching procedure selected 7,473 G6PD deficient subjects with 29,892 control subjects (4:1 control ratio) of the same gender, age, socioeconomic category, ethnic group, and year of first recorded visit in LHS (354 G6PD deficient individuals were left out for lack of proper matching controls). The demographic characteristics of the two groups are shown in Table 1. The mean patient age was 29.2 ± 22.3 years, 68.7% were men, and the mean follow-up time was 14.3 ± 6.2 years in both groups. There were no statistically significant differences in gender, age, ethnic and socio-economic groups and follow-up time between the two study groups.

**Table 1.** Demographic characteristics of G6PD deficient subjects and matched controls.

|  |  | **G6PD**<br>**Deficiency** | **Control** | **p** |
| --- | --- | --- | --- | --- |
| **N** |  | **7,473** | **29,892** |  |
| **Gender,**<br>**n (%)** | **Female** | 2,327 (31.1 %) | 9,308 (31.1 %) | 1.000 |
|  | **Male** | 5,146 (68.9 %) | 20,584 (68.9 %) | 1.000 |
| **Age (years)** | **mean ± SD** | 29.2 ± 22.3 | 29.2 ± 22.3 | 0.998 |
| **follow-up (years)** | **mean ± SD** | 14.3 ± 6.2 | 14.3 ± 6.2 | 0.293 |
| **age category,**<br>**n (%)** | **0- 2** | 378 (5.06 %) | 1,512 (5.06 %) | 1.000 |
|  | **3- 9** | 1,331 (17.81 %) | 5,324 (17.81 %) | 1.000 |
|  | **10-18** | 1,268 (16.97 %) | 5,072 (16.97 %) | 1.000 |
|  | **19-29** | 1,355 (18.13 %) | 5,420 (18.13 %) | 1.000 |
|  | **30-39** | 1,053 (14.09 %) | 4,212 (14.09 %) | 1.000 |
|  | **40-49** | 623 (8.34 %) | 2,488 (8.32 %) | 0.963 |
|  | **50-59** | 531 (7.11 %) | 2,128 (7.12 %) | 0.980 |
|  | **60-69** | 460 (6.16 %) | 1,841 (6.16 %) | 1.000 |
|  | **70-79** | 260 (3.48 %) | 1,039 (3.48 %) | 1.000 |
|  | **80-89** | 152 (2.03 %) | 608 (2.03 %) | 1.000 |
|  | **>= 90** | 62 (0.83 %) | 248 (0.83 %) | 1.000 |
| **Socio-economic**<br>**status (1-20)** | **mean ± SD** | 9.19 ± 3.56 | 9.14 ± 3.64 | 0.318 |
|  | **missing** | 708 (9.47 %) | 2,832 (9.47 %) |  |
| **Ethnic group,**<br>**n (%)** | **Arab** | 729 (9.76 %) | 2,916 (9.76 %) | 1.000 |
|  | **General** | 5,096 (68.19 %) | 20,384 (68.19 %) | 1.000 |
|  | **Jewish Ultra-orthodox** | 1,648 (22.05 %) | 6,592 (22.05 %) | 1.000 |
| **Smoking status, n**<br>**(%)** | **Non-smoker** | 3,505 (76.93 %) | 13,073 (74.77 %) | 0.002 |
|  | **Past smoker** | 83 (1.82 %) | 316 (1.81 %) | 0.950 |
|  | **Smoker** | 968 (21.25 %) | 4,096 (23.43 %) | 0.002 |
|  | **missing** | 2,917 (39.03 %) | 12,407 (41.51 %) |  |
| **BP systolic**<br>**(mmHg)** | **mean ± SD** | 119.8 ± 16.0 | 120.2 ± 16.3 | 0.228 |
|  | **missing** | 2,298 (30.8 %) | 9,568 (32.0 %) |  |
| **BP diastolic**<br>**(mmHg)** | **mean ± SD** | 71.8 ± 10.2 | 72.4 ± 10.4 | 0.001 |
|  | **missing** | 2,299 (30.8 %) | 9,568 (32.0 %) |  |
| **BMI**<br>**(kg/m2)** | **mean ± SD** | 23.3 ± 6.2 | 23.4 ± 6.4 | 0.104 |
|  | **missing** | 740 (9.90 %) | 3,626 (12.13 %) |  |
| **BMI**<br>**category** | **<18.5: Underweight** | 1,673 (24.8 %) | 6,590 (25.1 %) | 0.693 |
|  | **18.5-24.9: Normal** | 2,570 (38.2 %) | 9,837 (37.5 %) | 0.278 |
|  | **25-29.9: Overweight** | 1,575 (23.4 %) | 5,905 (22.5 %) | 0.114 |
|  | **>=30: Obese** | 915 (13.6 %) | 3,934 (15.0 %) | 0.004 |
|  | **missing** | 740 (9.9 %) | 3,626 (12.1 %) |  |

### Comorbidities of G6PD subjects and controls

The proportion of individuals in the two groups who had at least one diagnosis record for the studied conditions are presented in Table 2, together with the odds ratio (OR) and 95% confidence intervals (CI).

**Table 2.** Clinical characteristics of G6PD deficient subjects and matched controls.

|  | **G6PD**<br>**Deficiency** | **Control** | **p** | **Odds Ratio**<br>**[95% CI]** |
| --- | --- | --- | --- | --- |
| **N** | **7,473** | **29,892** |  |  |
| ***General comorbidities, n (%)*** |  |  |  |  |
| Arterial hypertension | 804 (10.8 %) | 3,365 (11.3 %) | 0.226 | 0.95 [0.87 to 1.03] |
| Diabetes mellitus | 418 (5.59 %) | 1,952 (6.53 %) | 0.003 | 0.85 [0.76 to 0.95] |
| Ischemic heart disease | 252 (3.37 %) | 909 (3.04 %) | 0.146 | 1.11 [0.96 to 1.28] |
| Chronic Obstructive Pulmonary Disease | 212 (2.84 %) | 738 (2.47 %) | 0.077 | 1.15 [0.98 to 1.35] |
| Chronic kidney disease | 109 (1.46 %) | 364 (1.22 %) | 0.105 | 1.20 [0.96 to 1.49] |
| ***Infectious diseases, n (%)*** |  |  |  |  |
| Acute sinusitis | 1,424 (19.1 %) | 5,091 (17.0 %) | <0.001 | 1.15 [1.07 to 1.22] |
| Acute bronchitis / bronchiolitis | 2,083 (27.9 %) | 8,442 (28.2 %) | 0.536 | 0.98 [0.93 to 1.04] |
| Pneumonia, bacterial | 275 (3.68 %) | 962 (3.22 %) | 0.047 | 1.15 [1.00 to 1.32] |
| Acute tonsillitis | 4,112 (55.0 %) | 15,411 (51.6 %) | <0.001 | 1.15 [1.09 to 1.21] |
| Cellulitis | 1,688 (22.6 %) | 6,038 (20.2 %) | <0.001 | 1.15 [1.08 to 1.23] |
| Acute gastroenteritis | 3,690 (49.4 %) | 13,865 (46.4 %) | <0.001 | 1.13 [1.07 to 1.19] |
| Urinary tract infection | 1,490 (19.9 %) | 5,182 (17.3 %) | <0.001 | 1.19 [1.11 to 1.27] |
| Viral infections (unspecified) | 5,032 (67.3%) | 18,968 (63.5%) | <0.001 | 1.19 [1.12 to 1.25] |
| Staphylococcal infections | 50 (0.67 %) | 124 (0.41 %) | 0.006 | 1.62 [1.14 to 2.27] |
| ***Atopic diseases, n (%)*** |  |  |  |  |
| Allergic rhinitis | 1,062 (14.2 %) | 3,732 (12.5 %) | <0.001 | 1.16 [1.08 to 1.25] |
| Allergic conjunctivitis | 859 (11.49 %) | 2,806 (9.39 %) | <0.001 | 1.25 [1.15 to 1.36] |
| Allergic urticaria | 722 (9.66 %) | 2,562 (8.57 %) | 0.003 | 1.14 [1.04 to 1.24] |
| Allergy to food/unspecified | 1,202 (16.1 %) | 4,031 (13.5 %) | <0.001 | 1.23 [1.15 to 1.32] |
| Asthma | 1,110 (14.9 %) | 4,166 (13.9 %) | 0.043 | 1.08 [1.00 to 1.16] |
| Atopic dermatitis | 1,110 (14.9 %) | 4,361 (14.6 %) | 0.571 | 1.02 [0.95 to 1.10] |
| Contact dermatitis | 1,632 (21.8 %) | 5,700 (19.1 %) | <0.001 | 1.19 [1.11 to 1.26] |
| ***Systemic autoimmune diseases, n (%)*** |  |  |  |  |
| Rheumatoid arthritis | 98 (1.31 %) | 164 (0.55 %) | <0.001 | 2.41 [1.85 to 3.12] |
| Systemic lupus erythematosus | 25 (0.335 %) | 22 (0.074 %) | <0.001 | 4.56 [2.46 to 8.48] |
| Sjogren's disease | 21 (0.28 %) | 30 (0.10 %) | 0.001 | 2.80 [1.53 to 5.07] |
| Scleroderma | 12 (0.161 %) | 7 (0.023 %) | <0.001 | 6.87 [2.49 to 20.59] |
| Dermatomyositis | 16 (0.214 %) | 18 (0.060 %) | <0.001 | 3.56 [1.70 to 7.40] |
| Ankylosing spondylitis | 22 (0.29 %) | 35 (0.12 %) | 0.001 | 2.52 [1.41 to 4.42] |
| ***Other inflammatory conditions, n (%)*** |  |  |  |  |
| Hashimoto's thyroiditis | 277 (3.71 %) | 889 (2.97 %) | 0.001 | 1.26 [1.09 to 1.44] |
| Graves' disease | 113 (1.51 %) | 312 (1.04 %) | 0.001 | 1.46 [1.16 to 1.81] |
| Pernicious anemia | 14 (0.187 %) | 3 (0.010 %) | <0.001 | 18.70 [5.22 to 101.6] |
| Idiopathic pulmonary fibrosis | 15 (0.201 %) | 25 (0.084 %) | 0.009 | 2.40 [1.18 to 4.74] |
| Hidradenitis suppurativa | 25 (0.33 %) | 39 (0.13 %) | <0.001 | 2.57 [1.49 to 4.36] |
| Fibromyalgia | 167 (2.23 %) | 342 (1.14 %) | <0.001 | 1.98 [1.63 to 2.39] |
| Psoriasis | 247 (3.31 %) | 836 (2.80 %) | 0.021 | 1.19 [1.02 to 1.37] |
| Inflammatory Bowel Disease | 60 (0.80 %) | 179 (0.60 %) | 0.051 | 1.34 [0.98 to 1.81] |
| Celiac disease | 33 (0.44 %) | 95 (0.32 %) | 0.120 | 1.39 [0.91 to 2.09] |
| ***Malignancies, n (%)*** |  |  |  |  |
| Hematologic malignancies | 42 (0.56 %) | 171 (0.57 %) | 1.000 | 0.98 [0.68 to 1.39] |
| Solid tumors | 224 (3.00 %) | 859 (2.87 %) | 0.563 | 1.04 [0.90 to 1.21] |

### General comorbidities

The proportion of patients with hypertension, ischemic heart disease, chronic obstructive pulmonary disease (COPD) and chronic kidney disease were not statistically different in the two groups, but there were comparably fewer individuals with diabetes mellitus in the G6PD deficient group (OR 0.85, 95% CI, 0.76 to 0.95, p=0.003).

### Infectious diseases

Most infectious diseases tested were significantly more frequent among G6PD deficient subjects than in controls (Table 2), with odds ratios typically around 1.15, with the notable exception of acute bronchitis/bronchiolitis which had a similar cumulative occurrence of ∼28% in the two groups. Strong associations were observed for urinary tract infections (OR 1.19, 95% CI 1.14 to 1.27, P<0.001) and unspecified viral infections (OR 1.19, 95% CI 1.12 to 1.25, P<0.001). Staphylococcal infections were particularly more frequent among G6PD deficient patients (OR 1.62, 95% CI 1.14 to 2.27, P<0.001).

### Allergic/Atopic conditions

For most of the atopic or allergic conditions assessed, we observed a higher prevalence among G6PD deficient individuals than in the control group (Table 2), notably for allergic conjunctivitis (OR 1.25, 95% CI 1.15 to 1.36, P<0.001), allergy to food/unspecified (OR 1.23, 95% CI 1.15 to 1.32, P<0.001), contact dermatitis (OR 1.19, 95% CI 1.15 to 1.32, P<0.001) and allergic urticaria (OR 1.14, 95% CI 1.04 to 1.24, P<0.001). Asthma was slightly more frequent among G6PD deficient individuals (OR 1.08, 95% CI 1.00 to 1.16, P=0.043). In contrast, atopic dermatitis rate was similar among G6PD deficient patients and the control group.

### Systemic autoimmune diseases

Systemic autoimmune diseases were strikingly more frequent in the G6PD group (Table 2). The association was particularly notable for scleroderma (OR 6.87, 95% CI 2.49 to 20.59, P<0.001), systemic lupus erythematosus (SLE) (OR 4.56, 95% CI 2.46 to 8.48, P<0.001), dermatomyositis (OR 3.56, 95% CI 1.70 to 7.40, P<0.001). Major risk associations were also observed for rheumatoid arthritis (OR 2.41, 95% CI 1.85 to 3.12, P<0.001), Sjögren’s syndrom (OR 2.80, 95% CI 1.53 to 5.07, P=0.001) and ankylosing spondylitis (OR 2.52, 95% CI 1.41 to 4.42, P=0.001).

We confirmed these finding by querying the proportion of individuals in each group who had purchased at least once, medications commonly used for autoimmune diseases, such as methotrexate (OR 2.30, 95% CI 1.74 to 3.02, P<0.001), hydroxychloroquine (OR 2.78, 95% CI 2.08 to 3.71, P<0.001), etanercept (OR 3.51, 95% CI 2.03 to 6.02, P<0.001) and leflunomide (OR 3.55, 95% CI 1.93 to 6.47, P<0.001). We also queried serological tests commonly used to diagnose autoimmune conditions (Table 3A). The G6PD deficient group had significantly higher rates of positive serological tests for antinuclear (OR 1.81), anti-histones (OR 2.96), RNP-68 (OR 4.00), and anti-Smith (OR 5.00) antibodies.

**Table 3.** Autoimmune serology in G6PD deficient subjects and matched controls.

|  | **G6PD**<br>**Deficiency** | **Control** | **p** | **Odds Ratio**<br>**[95% Confidence**<br>**Interval]** |
| --- | --- | --- | --- | --- |
| **N** | **7,122** | **28,488** |  |  |
| **A. Antinuclear Antibodies (Ab)** |  |  |  |  |
| Antinuclear Ab (EIA) positive | 246 (3.29 %) | 551 (1.84 %) | <0.001 | 1.81 [1.55 to 2.12] |
| Histones Auto-Ab positive | 17 (0.227 %) | 23 (0.077 %) | 0.001 | 2.96 [1.48 to 5.79] |
| RNP 68 Ab positive | 8 (0.107 %) | 8 (0.027 %) | 0.007 | 4.00 [1.31 to 12.24] |
| Smith (Sm) Ab positive | 5 (0.067 %) | 4 (0.013 %) | 0.020 | 5.00 [1.08 to 25.21] |
| **B. Cell-specific Antibodies (Ab)** |  |  |  |  |
| Anti Gastric Parietal Cell Ab positive | 16 (0.214 %) | 22 (0.074 %) | 0.002 | 2.91 [1.43 to 5.81] |
| Anti-Thyroid Peroxidase (TPO) positive | 82 (1.10 %) | 227 (0.76 %) | 0.005 | 1.45 [1.11 to 1.88] |

### Other inflammatory conditions

We further compared the two groups for a few organ specific inflammatory conditions. We observed significantly higher rate of Hashimoto’s thyroiditis (OR 1.26, 95% CI 1.09 to 1.44, P=0.001) and Graves’ disease (OR 1.46, 95% CI 1.16 to 1.81, P=0.03). Significantly higher rates were also observed for pernicious anemia (OR 18.70, 95% CI 5.22 to 101.6, P<0.001, idiopathic pulmonary fibrosis (OR 2.40, 95% CI 1.18 to 4.74, P=0.009) and hidradenitis suppurativa (OR 2.57, 95% CI 1.49 to 4.36, P<0.001). Psoriasis rate was also elevated to a lesser extent (OR 1.19, 95% CI 1.02 to 1.37, P=0.02). Interestingly fibromyalgia rate was significantly higher in the G6PD deficient group (OR 1.98, 95% CI 1.63 to 2.39, P<0.001). Inflammatory bowel disease (IBD) was more frequent with borderline statistical significance (OR=1.34, 95% CI 0.98 to 1.81, P=0.05). We confirmed the elevated rates of pernicious anemia and thyroiditis by comparing the rate of positive tests for anti-gastric parietal cells and anti-TPO in the two groups. Both were significantly elevated (Table 3B).

### Malignancy conditions

There were no statistically significant differences in the rates of hematologic malignancies and solid tumors.

### Laboratory tests in G6PD subjects and controls

To better understand the biological basis of these associations, we compared laboratory test results of individuals from the two groups. Table 4 displays the median and interquartile ranges (IQR) of the last test performed per patient together with the p-value for comparison and standardized mean difference (SMD).

**Table 4.** Laboratory characteristics of G6PD deficient subjects and matched controls.

|  | **G6PD Deficiency**<br>**(median [IQR])** | **Control**<br>**(median [IQR])** | **p** | **SMD** |
| --- | --- | --- | --- | --- |
| **Blood Count** |  |  |  |  |
| Red Blood Cells (M/uL) | 4.59 [4.26-4.92] | 4.83 [4.50-5.16] | <0.001 | **-0.419** |
| Hemoglobin (g/dL) | 13.00 [11.90-14.30] | 13.40 [12.20-14.70] | <0.001 | **-0.225** |
| Hematocrit (%) | 38.80 [35.70-42.50] | 39.60 [36.40-43.30] | <0.001 | **-0.165** |
| White Blood Cells (K/uL) | 7.01 [5.75-8.85] | 7.25 [5.94-9.07] | <0.001 | **-0.068** |
| Neutrophils (K/uL) | 3.41 [2.61-4.47] | 3.51 [2.68-4.60] | <0.001 | **-0.058** |
| Neutrophils (%) | 51.70 [42.20-59.40] | 51.20 [42.00-59.00] | 0.150 | **0.011** |
| Lymphocytes (K/uL) | 2.40 [1.87-3.23] | 2.47 [1.92-3.32] | <0.001 | **-0.047** |
| Lymphocytes (%) | 36.0 [29.0-44.5] | 36.2 [29.0-44.5] | 0.817 | **0.006** |
| Eosinophils (K/uL) | 0.19 [0.11-0.32] | 0.19 [0.12-0.33] | 0.002 | **-0.035** |
| Eosinophils (%) | 2.70 [1.70-4.40] | 2.70 [1.70-4.50] | 0.560 | **-0.008** |
| Basophils (K/uL) | 0.04 [0.03-0.06] | 0.04 [0.03-0.06] | 0.742 | **-0.007** |
| Basophils (%) | 0.60 [0.40-0.80] | 0.60 [0.40-0.80] | <0.001 | **0.053** |
| Monocytes (K/uL) | 0.54 [0.43-0.69] | 0.58 [0.46-0.73] | <0.001 | **-0.141** |
| Monocytes (%) | 7.80 [6.60-9.20] | 8.00 [6.80-9.50] | <0.001 | **-0.096** |
| Platelets (K/uL) | 265 [219-321] | 260 [214-318] | <0.001 | **0.059** |
| Mean Platelet Volume (fL) | 10.80 [10.20-11.60] | 10.80 [10.10-11.50] | <0.001 | **0.052** |
| **Biochemistry** |  |  |  |  |
| Fasting glucose (mg/dL) | 89.00 [83.4-96.0] | 89.6 [83.8-97.1] | <0.001 | **-0.041** |
| Creatinine (mg/dL) | 0.70 [0.52-0.85] | 0.71 [0.54-0.87] | <0.001 | **-0.050** |
| Creatine Kinase (U/L) | 87.0 [60.0-127.0] | 95.0 [66.0-138.0] | <0.001 | **-0.036** |
| Urea (mg/dL) | 27.2 [22.3-33.0] | 27.2 [22.2-33.2] | 0.524 | **0.005** |
| Uric Acid (mg/dL) | 4.93 [3.95-5.98] | 5.06 [4.04-6.11] | <0.001 | **-0.082** |
| Iron (ug/dL) | 94.0 [66.4-125.7] | 80.4 [58.5-105.8] | <0.001 | **0.369** |
| Ferritin (ng/mL) | 69.0 [33.8-144.3] | 48.0 [23.5-100.7] | <0.001 | **0.282** |
| Tranferrin (mg/dL) | 258 [233-288] | 266 [240-295] | <0.001 | **-0.149** |
| Tranferrin Saturation (%) | 26.2 [18.0-35.7] | 21.9 [15.6-29.0] | <0.001 | **0.401** |
| Bilirubin, Total (mg/dL) | 0.76 [0.55-1.09] | 0.56 [0.42-0.78] | <0.001 | **0.225** |
| Total Protein (g/dL) | 7.18 [6.89-7.47] | 7.11 [6.81-7.41] | <0.001 | **0.148** |
| Globulin (g/dL) | 2.86 [2.60-3.12] | 2.80 [2.56-3.06] | <0.001 | **0.129** |
| **Immunoglobulins** |  |  |  |  |
| Immunoglobulin A (mg/dL) | 186 [112-267.5] | 159 [97-229] | <0.001 | **0.256** |
| Immunoglobulin G (mg/dL) | 1135 [945.5-1323] | 1049 [883-1230] | <0.001 | **0.252** |
| Immunoglobulin M (mg/dL) | 99 [70-141] | 96 [68-135] | 0.069 | **0.049** |
| **Inflammatory Markers** | **(mean ± std. dev.)** | **(mean ± std. dev.)** |  |  |
| Erythrocyte sedimentation rate (mm/hr) | 20.69 ± 19.82 | 18.98 ± 18.14 | <0.001 | **0.093** |
| C-reactive protein (mg/L) | 7.72 ± 19.69 | 7.86 ± 20.58 | 0.147 | **-0.007** |
| C3 Complement (mg/dL) | 128.38 ± 24.55 | 130.44 ± 23.71 | 0.038 | **-0.086** |
| C4 Complement (mg/dL) | 31.71 ± 10.46 | 33.52 ± 10.63 | <0.001 | **-0.170** |

IQR: Interquartile range
SMD: Standardized Mean Difference

As expected for a condition associated with hemolysis, we observe significantly lower levels of red blood cells (P<0.001, SMD=-0.419), hemoglobin (P<0.001, SMD=-0.225) and hematocrit (P<0.001, SMD=-0.165) in the G6PD deficient group. Interestingly, we also observe lower levels of white blood cells (WBC) (P<0.001, SMD=-0.068) in general, and more specifically, diminished monocytes (P<0.001, SMD=-0.141) and neutrophils (P<0.001, SMD=-0.058). The breakdown of cell types among WBC is not significantly different in G6PD, except for monocytes, which have a decreased percentage among WBC (P<0.001, SMD=-0.096), whereas the percentage of basophils is increased in G6PD deficient individuals (P<0.001, SMD=0.053). Platelets are also present in higher numbers in G6PD deficient patients (P<0.001, SMD=0.059) with increased mean platelet volume (P<0.001, SMD=0.052).

In biochemistry tests, we observe lower fasting glucose, creatinine and creatine kinase levels among G6PD deficient patients. Conspicuous differences are observed for iron, ferritin and transferrin saturation, which are elevated with respective SMDs of 0.369, 0.282 and 0.401 (P<0.001 for all), while transferrin is markedly decreased (SMD=-0.149). Interestingly both blood total protein and globulin are markedly elevated in G6PD deficiency, with respective SMDs of 0.148 and 0.129 (P<0.001 for both). Among immunoglobulins (Ig), we observe markedly elevated levels for IgA and IgG with respective SMDs of 0.256 and 0.252 (P<0.001 for both). In contrast, IgM levels are not significantly different in the two groups.

We also looked at inflammatory markers. Erythrocyte sedimentation rate (ESR) is slightly higher in G6PD patients (P<0.001, SMD=0.093), but not C-reactive protein. Levels of C4 complement are significantly decreased in G6PD deficient patients (P<0.001, SMD=-0.170), but C3 complement are only slightly decreased (P=0.038, SMD=-0.086).

## Discussion

To our knowledge, this is the first large-scale epidemiologic analysis to assess the association between G6PD deficiency and the long-term risk of immune system associated diseases. We show G6PD deficient patients have globally higher rates of acute infectious diseases, and in particular staphylococcal infections, suggesting a predisposition to infections. The link between G6PD deficiency and infections has been previously recognized in infancy, with lack of G6PD activity identified as a risk factor for neonatal sepsis in males (Rostami-Far et al., 2016). This study is the first to show this association on a large-scale and for a wide range of infectious diseases.

Another finding from our study is a higher rate of allergic diseases in G6PD-deficient subjects. No systematic review of allergic comorbidity in G6PD deficiency was found in the literature. However, a retrospective case-control study showed that G6PD deficiency is an independent risk factor for asthma (Fois et al., 2021) and G6PD was noticeably downregulated in asthmatic children (Li et al., 2020). In asthma, the level of reduced GSH is significantly increased, and G6PD-deficient subjects’ insufficient production of NADPH may contribute to asthma pathogenesis (Husain et al., 2012). Moreover, the chronic depletion of nitric oxide in G6PD deficient subjects may affect the basal bronchodilator tone (Prado et al., 2011). In our cohort, we also found an association between asthma and G6PD deficiency (OR=1.08, P=0.043), but much more stronger associations were present for other conditions such as allergic conjunctivitis (OR=1.25, P<0.001), food allergy (OR=1.23, P<0.001), and contact dermatitis (OR=1.19, P<0.001).

The most striking finding of our study are the much-elevated rates of autoimmune diseases and chronic inflammatory conditions in G6PD deficient individuals. We notably observed markedly elevated rates of scleroderma (OR=6.87, P<0.001), SLE (OR=4.56, P<0.001), dermatomyositis (OR=3.56, P<0.001), Sjögren’s syndrome (OR=2.80), rheumatoid arthritis (OR=2.41, P<0.001) and ankylosing spondylitis (OR=2.52, P=0.001) in individuals affected with G6PD deficiency. These findings were corroborated with elevated rates of positive autoimmune serology in G6PD deficiency and higher rates of treatment with medications commonly used to treat autoimmune conditions. This is consistent with a previous study reporting a lower level of G6PD enzymatic activity in patients with rheumatoid arthritis and Sjögren’s syndrome (Gheita et al., 2014).

We also observed significantly higher rates of organ-specific inflammatory diseases such as Hashimoto’s thyroiditis (OR=1.26, P=0.001) and Graves’ disease (OR=1.46, P=0.001), and particularly elevated rates of pernicious anemia (OR=18.7, P<0.001). We corroborated these findings with higher rates of individuals with positive anti-gastric parietal cells and anti-TPO antibodies in the G6PD group. Findings of increased rate of autoimmune thyroid disorders in G6PD patients are consistent with a recently published case-control study from North Sardinia, who identified an odds ratio of 1.36 (95% CI 1.11 to 1.60) for these disorders for G6PD patients (Dore et al., 2023).

We also observed higher rates of inflammatory bowel disease (OR=1.34, P=0.05) and psoriasis (OR=1.19, P=0.02) among G6PD deficient patients. Interestingly, we observed markedly higher rates for diseases for which the pathological basis is poorly understood, notably hidradenitis suppurativa (OR=2.57, P<0.001) and fibromyalgia (OR=1.98, P<0.001). These strong associations with G6PD deficiency suggest that metabolic pathway alterations, as those present in G6PD deficiency, may play a major role in these pathologies.

G6PD deficient individuals differed markedly from the rest of the population for commonly prescribed laboratory tests. They display diminished white blood cells counts, particularly monocytes and neutrophils. Interestingly, activated monocytes and neutrophils produce reactive oxygen species (ROS), which could be particularly harmful to G6PD-deficient cells and could lead to increased cell death. We also observed markedly increased levels of iron, ferritin, and transferrin saturation, with diminished transferrin levels. Immunoglobulins A and G levels were increased in G6PD patients and C4 complement deceased, suggesting a possible activation of the complement system in G6PD deficiency, presumably through the classical pathway and with the production of circulating immune complexes (Wang and Liu, 2021). Indeed, immune dysregulation may play a role in the pathogenesis of autoimmune and infectious diseases in G6PD deficiency, presumably through dysregulated NADPH homeostasis and ROS imbalances (Brewer et al., 2013).

G6PD deficiency has been associated with a proinflammatory state with over-expression of IL-8, IL-4, IL-5, and IL-9 cytokines, and increased chemotaxis of eosinophils and Th2 immune polarization (Parsanathan and Jain, 2021; Yang et al., 2015). Oxidative stress had been suggested in the pathogenesis of human SLE and Sjögren syndrome (Kurien et al., 2011; Neumann et al., 2003). Oxidative stress in G6PD deficient subjects contributes to developing novel epitopes by oxidizing proteins and lipids and producing anti-DNA antibodies (Lee, 2003; Zhang et al., 2011). Oxidative stress may also play a role in the sustained activation of lymphocytes and cause autoimmune responses by modulation of T and B cell homeostasis, leading to a loss in immunological tolerance (Piera-Velazquez and Jimenez, 2019; Yeh et al., 2013). These may be some of the reasons for the observed higher rates of autoimmune diseases. Their roles in food allergy, atopic and allergic contact dermatitis should be elucidated in further research.

Autoimmune diseases in G6PD deficient patients may be triggered by activation of TGF-β/NADPH oxidases/ROS signaling, the expression of ICAM- 1 and VCAM-1, and the adhesion of leukocytes to the endothelial cells with endothelial to mesenchymal transition (Hal Scofield et al., 2005; Parsanathan and Jain, 2021).

Several mechanisms could explain higher susceptibility to infections in individuals with G6PD deficiency. NADPH is critical for the regeneration of glutathione (GSH), reactive oxygen species (ROS) and plays an essential role in the production of reactive nitrogen species (RNS) and nitric oxide (NO) (Yang et al., 2011). Immune inflammation in airway epithelial cells induces G6PD activity with the concomitant increase of GHS, ROS, nitrotyrosine, and NADPH oxidase 2 (NOX2) (Nadeem et al., 2018). Experimental models have shown that G6PD inhibition suppresses airway inflammation induced by lipopolysaccharides and the ROS derived from NOX2 (Hsieh et al., 2013).

Dysregulated redox systems and NF-κB signaling in G6PD-deficient cells have been associated with increased susceptibility to coronavirus and enterovirus infection (Ho et al., 2008; Wu et al., 2015). G6PD-deficient individuals also produce lower levels of pro-inflammatory cytokines, IL-6, and IL-1β in peripheral mononuclear cells (Sanna et al., 2007), and their granulocytes display diminished bactericidal activity and increased susceptibility to infection (van Bruggen, 2002). Reduced expression of prostaglandin E2 and cyclooxygenase-2 (Lin et al., 2016), defective neutrophil extracellular trap (NET) formation (Cheng et al., 2013), and impaired inflammasome activation have also been observed in the neutrophils of G6PD-deficient individuals (Yen et al., 2020).

The lack of an association between G6PD deficiency and malignancy in our study is an important finding. Earlier studies with smaller sample sizes have observed decreased cancer risk in G6PD deficient patients, but many studies failed to replicate this finding. Here, in a large cohort of G6PD deficient individuals, carefully matched by multiple factors, we did not find evidence for an association between G6PD deficiency and the overall rate of both solid and hematologic cancers during a mean follow-up of 14.3 years. It is remarkable that even though G6PD deficiency is associated with several chronic inflammatory diseases, this does not translate into cancer risk. This may be explained by the fact that G6PD deficiency is associated with multiple changes in metabolic and cellular states (Beutler, 1994) with some changes being pro-tumorigenic and other changes being anti-tumorigenic.

This study has the usual limitations of observational studies: potential confounders could be distributed differently between the two groups, clinical records may be inaccurate, and data may be missing. Nonetheless, the large number of participants and the scrupulous matching methodology adopted should minimize the extent of such biases. Another limitation is that the study reflects the link with the G6PD mutations prevalent in the Israeli population, in which the most prevalent mutation is the “Mediterranean” allele, which is associated severe G6PD enzymatic deficiency (Kurdi-Haidar et al., 1990). The risks may differ with variants present in other populations. Since we lack G6PD enzymatic measures for the whole cohort, we considered G6PD deficiency as a dichotomous trait rather than a continuous spectrum.

In conclusion, G6PD deficiency is a condition which appears to be associated with increased susceptibility to autoimmune, infectious and allergic diseases. Additional studies are warranted to confirm these observations and investigate the mechanisms underlying the higher prevalence of these conditions in G6PD deficiency. Some of the novel associations we report with G6PD deficiency are for poorly understood conditions such as fibromyalgia or hidradenitis suppurativa, and this may suggest new directions for investigating the pathological basis of these diseases.

## Data Availability

Data were obtained from patients' electronic health records and IRB approval restrains its use to researchers inside Leumit Health Services.

## Acknowledgements

This research was supported in part by the Intramural Research Program, National Institutes of Health, National Cancer Institute, Center for Cancer Research. The content of this publication does not necessarily reflect the views or policies of the Department of Health and Human Services, nor does mention of trade names, commercial products, or organizations imply endorsement by the U.S. Government.

